# The Predictive Value of Diaphragm Muscle Ultrasound for Ventilator Weaning Outcomes after Cervical Spinal Cord Injury: A Retrospective Case Series

**DOI:** 10.1101/2024.02.28.24303260

**Authors:** Natasha S. Bhatia, Stephany Kunzweiler, Christopher Conley, Ki H. Kim, Adenike A. Adewuyi, Antonio Mondriguez-Gonzalez, Lisa F. Wolfe, Mary Kwasny, Colin K. Franz

**Affiliations:** Shirley Ryan AbilityLab, Chicago, IL, USA; Department of Physical Medicine and Rehabilitation, Northwestern University Feinberg School of Medicine, Chicago, IL, USA; Division of Pulmonary and Critical Care Medicine, Department of Medicine, Northwestern University Feinberg School of Medicine. Chicago, IL, USA; Ken and Ruth Davee Department of Neurology, Northwestern University Feinberg School of Medicine, Chicago, IL, USA; Department of Preventive Medicine, Northwestern University Feinberg School of Medicine, Chicago, IL, USA

**Keywords:** diaphragm muscle, neuromuscular, ultrasound, ventilator weaning, spinal cord injury

## Abstract

**Objectives:** Neuromuscular respiratory failure after cervical spinal cord injury (cSCI) can lead to dependence on an invasive mechanical ventilator. Ventilator-free breathing after cSCI is associated with improved morbidity, mortality, and quality of life. We investigated the use of diaphragm muscle ultrasound to predict ventilator weaning outcomes after cSCI.

**Methods:** This is a retrospective case series conducted at a university-affiliated freestanding inpatient rehabilitation facility. We identified patients with cSCI who had a tracheostomy and were dependent on an invasive mechanical ventilator at the time of admission to inpatient rehabilitation. A diaphragm muscle ultrasound was performed, which included measurements of the thickness of the diaphragm and a calculation of the thickening ratio (TR), which reflects diaphragm muscle contraction. The primary outcome measure was the need for mechanical ventilation at time of discharge from the inpatient rehabilitation facility. Successful ventilator weaning was defined as either daytime or full 24-hour ventilator-free breathing.

**Results:** Of the 21 patients enrolled, 11 (52%) were able to wean successfully (partially or fully) from the ventilator. Of the ultrasound measurements that were taken, the TR was the optimal predictor for ventilator weaning outcomes. A threshold of TR≥1.2 as the maximum hemidiaphragm measurement had a sensitivity of 1.0 and specificity of 0.90 for predicting ventilator weaning.

**Conclusion:** Normal diaphragm contractility (TR ≥ 1.2) as determined by diaphragm muscle ultrasound is a strong positive predictor for successful ventilator weaning in patients with cSCI.

Utilizing diaphragm ultrasound, rehabilitation physicians can set precision rehabilitation goals regarding ventilator weaning for inpatients with respiratory failure after cSCI, potentially improving both outcomes and quality of life.

## 1. Introduction

Dependence on a ventilator after cervical spinal cord injury (cSCI) has important implications for morbidity, mortality, and quality of life. People with cSCI who rely on mechanical ventilation are almost 40 times more likely to die during the first year after injury compared to those who are not ventilator-dependent,^1^ and long-term mortality rates remain up to 3.5 times higher for those who require mechanical ventilation after the first year of injury.^1,2^ This excess mortality is predominantly a result of respiratory disorders, and the most common causes of death for ventilator-dependent people with SCI are pneumonia and other respiratory complications.^3^ People with SCI who are ventilator-dependent report a more negative perception of their health status, lower social integration scores, and worse quality of life outcomes compared to people living with SCI who do not require a ventilator.^4^

Respiratory failure after spinal cord injury is a result of dysfunction of both ventilation and airway clearance.^5^ The neurologic level of injury (NLI) after cSCI is a critical factor in the need for mechanical ventilation. The primary muscle of inspiration, the diaphragm, is innervated by the phrenic nerve which originates from the C3, C4, and C5 nerve roots, with C4 providing primary innervation.^6^ Patients with a neurologically complete SCI at the C1-2 levels are typically not able to achieve ventilator-free breathing, though some studies have shown the possibility of weaning in rare cases.^7^ For those with a NLI at C3, ventilator-free breathing rates vary from 25- 50%.^7,8^ For those with NLI at C4, up to 70-80% of patients can successfully wean from the ventilator, and an even higher percentage achieve ventilator-free breathing with a NLI at C5-6 and below.^7–9^ This variability even within each neurologic level illustrates the need for additional clinical tools to both determine readiness for ventilator weaning and serve as an accurate predictor for successful ventilator-free breathing outcomes.

Current clinical tools used to predict successful ventilator-free breathing include bedside spirometry and respiratory function parameters (such as vital capacity and negative inspiratory force), fluoroscopic evaluation of the diaphragm, phrenic nerve conduction studies (NCS), and needle electromyography (EMG) of the diaphragm muscle.^9^ While bedside spirometry and fluoroscopic evaluation of the diaphragm are non-invasive methods to diagnose diaphragm dysfunction, they are less sensitive than other techniques.^9^ Conversely, while needle EMG and phrenic NCS are sensitive tests for diaphragm dysfunction, these methods are more invasive and require a highly skilled practitioner to perform the procedures and interpret the results.

Moreover, needle EMG of the diaphragm can be associated with risk of diaphragm injury and pneumothorax, while phrenic NCS has a high false positive rate associated with lack of response on NCS despite intact diaphragm muscle innervation.^10^

Ultrasound of the diaphragm muscle has been used to evaluate diaphragm function and contractility in the context of a variety of disorders including phrenic nerve injury, direct diaphragm injury, lung transplantation, and more recently, respiratory dysfunction after COVID- 19 infection.^11,12^ Prior work from Boon and colleagues has reported a 93% sensitivity and 100% specificity of this mode of diaphragm ultrasound in identifying neuromuscular diaphragm dysfunction from phrenic neuropathy.^11^

The purpose of this study was to investigate the efficacy of diaphragm muscle ultrasound to predict ventilator weaning outcomes after cSCI in patients dependent on mechanical ventilation.

## 2. Materials and Methods

This study is a retrospective case series via chart review of data obtained as part of routine patient care where the sonographer was not blinded to the patient’s diagnosis. Northwestern University’s Institutional Review Board (IRB) approved the study (STU23625789). This included a waiver of informed consent, justified by the minimal risk to subjects and the impracticality of obtaining consent for this review. The IRB’s approval ensured compliance with ethical principles, including patient privacy and data confidentiality, in line with the Declaration of Helsinki.

This study was conducted at a university-affiliated freestanding acute rehabilitation hospital. We identified patients with cSCI who had a tracheostomy and were dependent on invasive mechanical ventilation for any number of hours at the time of admission to the rehabilitation unit, and who were referred for a neuromuscular diaphragm ultrasound. As part of our institution’s interdisciplinary ventilator liberation program, all patients with cSCI on a ventilator who have a goal of potential ventilator weaning are referred for a diaphragm ultrasound evaluation. We collected demographic and clinical data including time from injury, American Spinal Injury Association (ASIA) Impairment Scale (AIS) neurologic level of injury and completeness of injury, and number of hours per day spent on the ventilator.

A practicing physician (CKF) board certified in both Physical Medicine and Rehabilitation and Neuromuscular Medicine (American Board of Physical Medicine and Rehabilitation), and who holds the Neuromuscular Ultrasound Certificate of Added Qualification (American Board of Electrodiagnostic Medicine), performed the diaphragm muscle ultrasound examinations. This study followed a protocol for diaphragm ultrasound evaluation as previously published by Farr et al, based on Boon’s original study, since it corresponds to the normative data set, and was taken from a similar population of healthy controls within the same geographic area as our study population.^10,13^ Typically, the sonographic evaluation includes a B-mode examination of each hemidiaphragm, with measurements of the thickness of the hemidiaphragm muscle both at maximal inspiration (i.e. total lung capacity, TLC) and at end expiration (i.e. functional residual capacity, FRC).^11^ At FRC in the zone of apposition, a diaphragm thickness of ≥0.15cm is considered normal, with a thickness less than this cutoff consistent with diaphragm atrophy.^10^ From these two measurements of diaphragm size, a thickening ratio (TR) can be calculated, where TR = thickness at TLC / thickness at FRC (**Figure 1**). The TR reflects diaphragm muscle contractility, with a normal TR≥1.2.^10^ Our sonographic evaluation included examination of each hemidiaphragm using a low intercostal approach (8^th^ or 9^th^ intercostal space) along the anterior axillary line **(Figure 2**). The ultrasound evaluation included measurements of the thickness of each hemidiaphragm at maximal inhalation (or TLC) and exhalation (or FRC), as well as a calculation of the TR as a marker of diaphragm contractility. These measures are taken bilaterally (one measure for each hemidiaphragm). In general, patients with only one paretic hemi-diaphragm are not expected to be completely ventilator dependent.^14^ As such, we preferred to use the maximum measurement of the two sides to determine if this function may predict the ability to wean from the ventilator.

**Figure 1.**
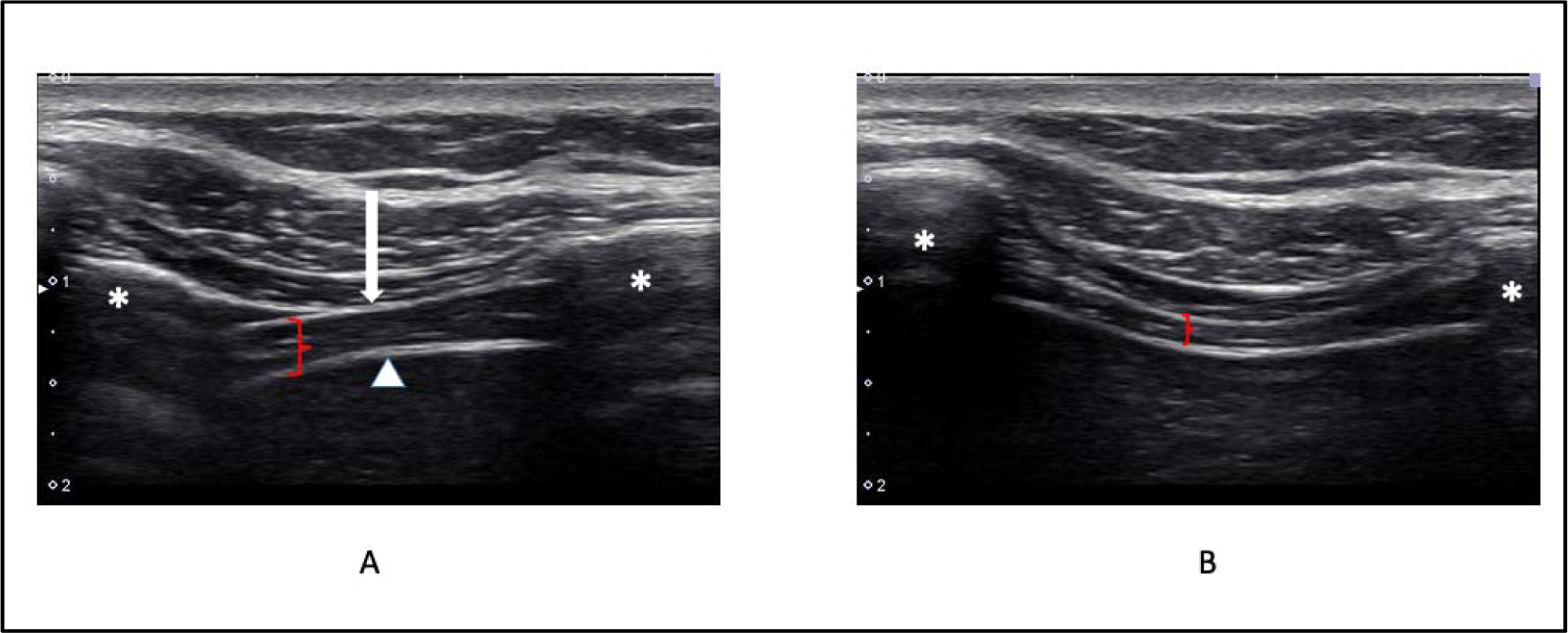
Diaphragm ultrasound images showing the diaphragm muscle thickening that occurs during inspiration and diaphragm contraction. (A) Ultrasound image of the diaphragm illustrating the thickness of the diaphragm (red bracket) at TLC, or inhalation. Asterisk, ribs. Arrow, pleural line. Arrow head, peritoneal line. (B) Ultrasound image of the diaphragm illustrating the thickness of the diaphragm (red bracket) at FRC. The thickening ratio is calculated as TR = thickness at TLC / thickness at FRC.

**Figure 2.**
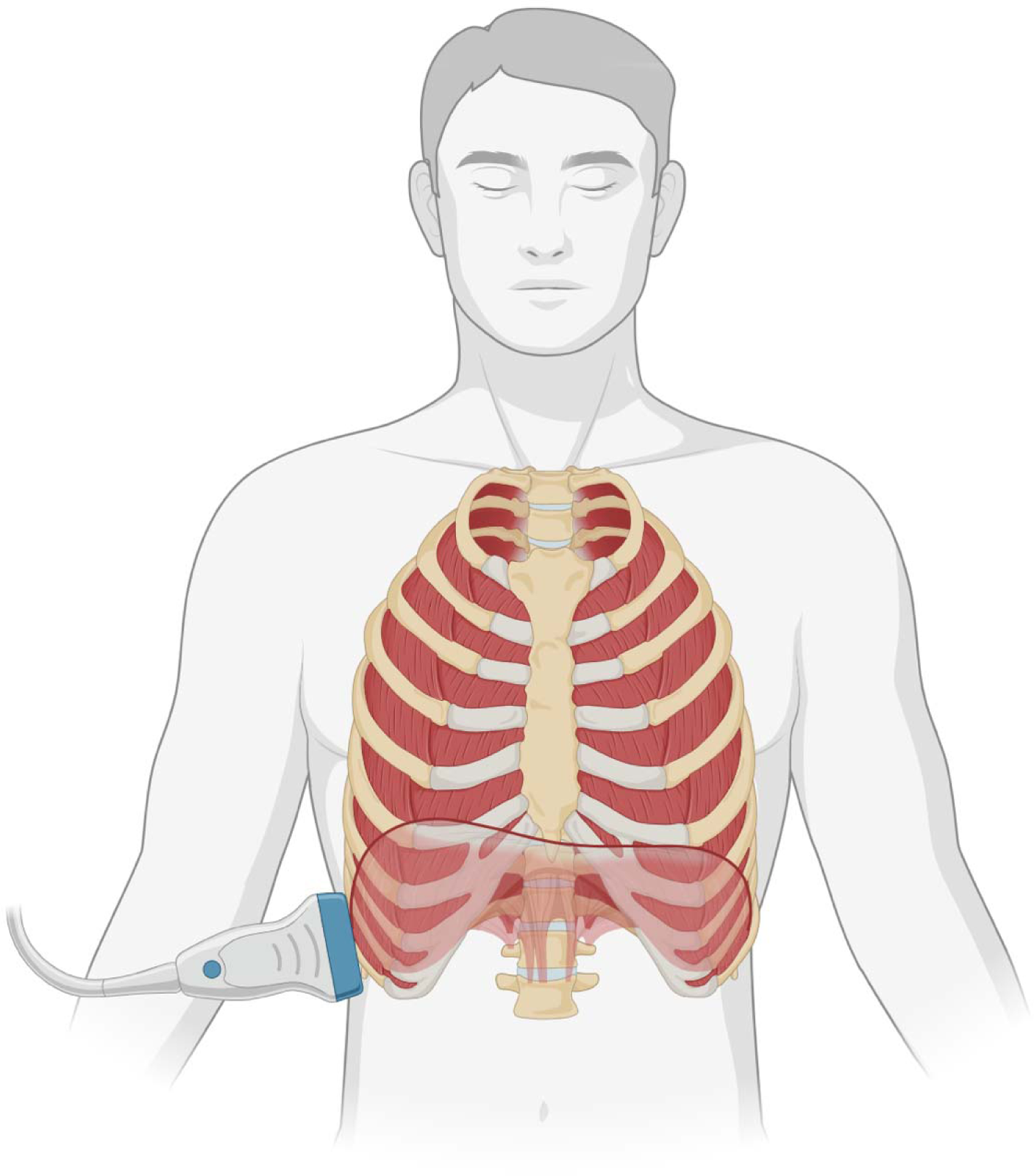
This figure demonstrates the positioning of an ultrasound transducer used in our protocol based on the technique described by Boon et al.^10^ The transducer is placed on a lower intercostal space (8th or 9th) along the anterior axillary line. *Figure created with BioRender.com*.

After sonographic evaluation, the medical team performed a clinical assessment of respiratory parameters (including vital capacity, negative inspiratory force, resting tidal volume, and CO2/arterial blood gas results) to determine readiness for ventilator weaning. Once these respiratory parameters fall within safe guidelines, the ventilator weaning process follows a progressive ventilator free breathing protocol, where patients spend progressively increasing intervals of time off the ventilator each day, and which is the most successful method of ventilator weaning for people with SCI (see **Supplemental File 1**).^15^

The primary outcome measure was the need for mechanical ventilation at the time of discharge from acute inpatient rehabilitation. The outcomes of patients in our data set could be grouped into two categories: no meaningful ventilator weaning (off ventilator for <1 hour per day), and successful ventilator weaning (including both daytime ventilator-free breathing with nocturnal use of the ventilator, and full ventilator weaning). Because daytime ventilator-free breathing is associated with improved outcomes and quality of life, analysis was performed by categorizing patients as either successfully weaned from the ventilator (full or daytime) or not weaned.

Logistic regression models were fit, and the area under the receiver operating characteristic curve was used to determine the best predictor of successful (full or partial) weaning. Sensitivity and specificity were then calculated.

## 3. Results

This retrospective case series followed 21 patients who were screened by ultrasound for their candidacy for ventilator weaning during their inpatient rehabilitation admission (**Table 1**). There were 19 male and 2 female patients, with an average age of 44 years (range = 17-78 years). All patients sustained a traumatic cSCI, ranging from C1-C5 neurologic level of injury, and AIS A-C completeness. All patients required mechanical ventilation via tracheostomy at the time of admission to acute inpatient rehabilitation.

**Table 1.** Demographic characteristics and weaning outcomes of patients undergoing diaphragm ultrasound evaluation, grouped by outcome.

| Patient # | Age/Sex | Neurologic Level | AIS | Thickening Ratio (Left) | Thickening Ratio (Right) | Time from Injury on Admission (Days) | Length of Weaning Process (Days) | Outcome at DC |
| --- | --- | --- | --- | --- | --- | --- | --- | --- |
| 1 | 70s/F | C4 | A | 1.4 | 2.5 | 25 | 10 | weaned |
| 2 | 40s/M | C4 | A | 1.4 | 1.1 | 25 | 11 | weaned |
| 3 | 60s/M | C4 | A | 1.5 | 2 | 10 | 90 | weaned |
| 4 | 30s/M | C4 | A | 1.4 | 1 | 22 | 18 | weaned |
| 5 | 20s/M | C4 | C | 1.2 | 1.1 | 46 | 32 | weaned |
| 6 | 60s/M | C5 | A | 1.4 | 1.2 | 40 | 27 | weaned |
| 7 | 60s/M | C4 | A | 1.2 | 1.4 | 36 | 51 | weaned |
| 8 | 20s/M | C5 | A | 1.5 | 2 | 60 | 10 | weaned |
| 9 | 40s/M | C2 | A | 1.3 | 1.3 | 35 | 64 | weaned |
| 10 | 20s/M | C5 | B | 1.7 | 1.3 | 41 | 17 | weaned |
| 11 | 20s/M | C2 | A | 1.7 | 1.5 | 30 | 93 | weaned |
| 12 | <20/M | C1 | A | 1 | 1.8 | 30 | no wean | no wean |
| 13 | 30s/F | C2 | A | 1 | 1 | 20 | no wean | no wean |
| 14 | 30s/M | C1 | C | 1 | 1 | 22 | no wean | no wean |
| 15 | 50s/M | C4 | A | 1 | 1 | 124 | no wean | no wean |
| 16 | 70s/M | C1 | C | 1.1 | 1.1 | 81 | no wean | no wean |
| 17 | 50s/M | C2 | C | 1 | 1 | 36 | no wean | no wean |
| 18 | 60s/M | C2 | A | 1 | 1 | 30 | no wean | no wean |
| 19 | 60s/M | C3 | A | 1 | 1 | 103 | no wean | no wean |
| 20 | <20/M | C1 | A | 1 | 1 | 32 | no wean | no wean |
| 21 | <20/M | C2 | B | 1 | 1 | 59 | no wean | no wean |

At time of discharge, 11 patients (52%) achieved successful (full or daytime) ventilator-free breathing, while 10 patients did not achieve meaningful ventilator-free breathing. The mean TR calculated from the best hemidiaphragm measurement for patients who achieved ventilator-free breathing was 1.6, while the mean TR for patients who did not achieve ventilator weaning was 1.1 (**Table 2**). Median TR was also calculated for both groups given outlier data within the data set. The successfully weaned group had a median TR of 1.4, and the unsuccessfully weaned group had a median TR of 1.0 (**Table 2**). Using a cutoff of TR ≥ 1.2 resulted in 100% sensitivity and 90% specificity for predicting successful ventilator weaning (**Table 3**).

**Table 2.**
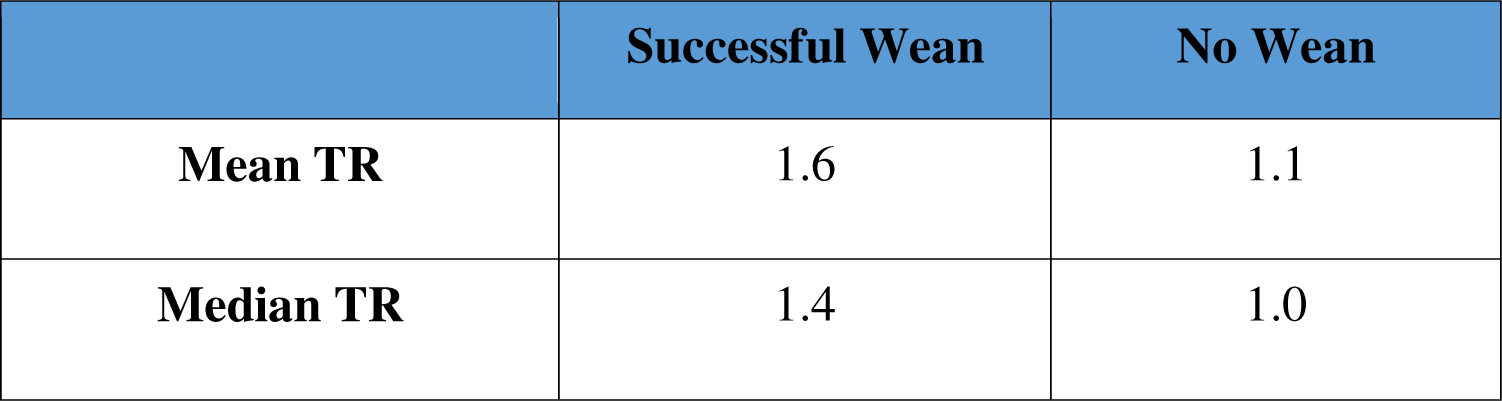
Mean and median diaphragm thickening ratios, calculated using each patient’s maximum hemidiaphragm thickening ratio if patient had asymmetrical diaphragm function.

**Table 3.** Sensitivity, specificity, positive predictive value, and negative predictive value using a TR≥ 1.2 to predict successful ventilator weaning.

|  | Successful Wean (n) | No Wean (n) |  |
| --- | --- | --- | --- |
| Maximum $TR \geq 1.2$ | 11 | 1 | PPV = 92% |
| Maximum $TR < 1.2$ | 0 | 9 | NPV = 100% |
|  | Sensitivity = 100% | Specificity = 90% |  |

Within our study population, all 11 patients who achieved successful ventilator weaning had a maximum TR of 1.2 or greater (**Table 3**), and 8 of those patients (72%) in this group had normal TRs bilaterally. Of the 10 patients who did not wean from the ventilator, 9 had impaired or absent contractility bilaterally, and only one patient had a normal maximum TR (patient #12), though did have their rehabilitation course complicated by recurrent pneumonia.

The receiver operating characteristic curve was plotted with an area under the curve of 0.95, which indicates a high overall performance of the best thickening ratio as a diagnostic test for successful ventilator weaning at the chosen cutoff of 1.2. The sensitivity at this cutoff was 100%, indicating that all successful weaning cases were correctly identified using this cutoff. The specificity was 90%, indicating that 90% of the failures were correctly identified as such when using the cutoff of 1.2. A high sensitivity and specificity suggest that a cutoff of 1.2 for the best thickening ratio is a good threshold for predicting successful ventilator weaning.

## 4. Discussion

The use of diaphragm ultrasound provides valuable information about the diaphragm muscle’s innervation status,^16^ which is especially important in the case of patients with C3 or C4 level injuries where diaphragm dysfunction may be a consequence of mixed upper and lower motor neuron lesions, so retained partial innervation cannot be assumed, and ventilator weaning outcomes are variable. The inability to accurately predict short- and long-term ventilator needs interferes with delivering optimal care for patients after cSCI.

Our findings demonstrate that a normal thickening ratio as determined by diaphragm ultrasound evaluation is a strong predictor of successful ventilator-free breathing. Evidence of intact contractility on ultrasound can indicate weaning potential for patients who would typically not be expected to wean from the ventilator based on their injury levels. In our study, for example, two patients with C2 AIS A injuries were found to have intact contractility bilaterally and were both able to successfully wean from the ventilator. Conversely, evidence of abnormal diaphragm contractility can also indicate the need for caution with ventilator weaning attempts. Two patients who would be expected to retain some degree of diaphragm innervation given their injury level (C3 and C4 levels of injury) both displayed absent contractility bilaterally and were ultimately unable to wean from the ventilator.

For patients unable to fully wean from the ventilator at time of discharge from acute care, options for ongoing rehabilitation are often limited. Many acute inpatient rehabilitation facilities cannot accept patients requiring mechanical ventilation due to inadequate staffing. In these cases, patients who require mechanical ventilation at time of discharge from acute care are sent to long-term acute care facilities to attempt to wean off the ventilator prior to admission to an inpatient rehabilitation unit. This results in a delay in intensive rehabilitation therapies, which are critical in the early stages of recovery from SCI. Acute inpatient rehabilitation facilities that can manage patients on a ventilator are thus uniquely able to both provide intensive rehabilitation therapies and potentially work towards ventilator weaning. Clinical tools that can help better identify which patients may be able wean from a ventilator successfully can help guide clinical practice and rehabilitation goal setting.

Within our treatment center, the application of our findings has contributed to the development of a treatment algorithm to guide our approach to ventilator weaning for patients with SCI (**Figure 3**). Specifically, our approach includes an evaluation of diaphragm atrophy and contractility during initial evaluation for patients with cSCI who are ventilator-dependent on admission.

**Figure 3.**
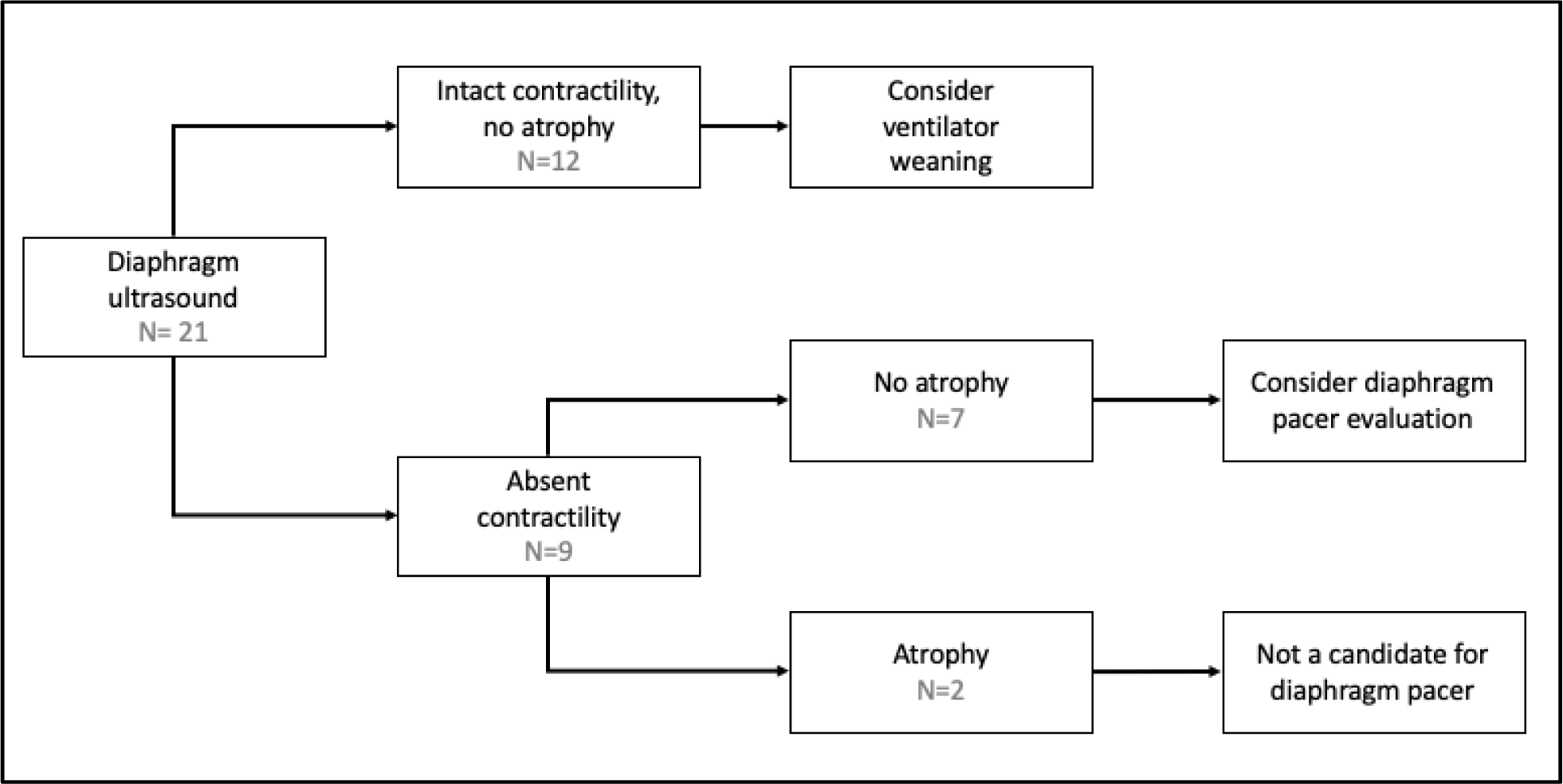
Clinical algorithm for ventilator weaning based on measurements of contractility and muscle thickness on diaphragm muscle ultrasound evaluation. Of the 21 participants in our study, 12 demonstrated intact contractility without significant atrophy, and were considered for ventilator weaning. Of the 9 participants with absent contractility, 7 showed normal diaphragm muscle thickness (no atrophy), and 2 of these eventually underwent diaphragm pacer placement. Of the patients with absent contractility, 2 also demonstrated significant atrophy, and were not candidates for diaphragm pacing.

Patients who demonstrate normal contractility of the diaphragm and no significant atrophy are deemed to be good candidates for eventual ventilator weaning, even when other traditional markers for readiness to wean, such as vital capacity or negative inspiratory force, are not indicative of successful weaning at the time of evaluation. For those with present but impaired contractility of the diaphragm, it is felt they may eventually be able to achieve some degree of ventilator-free breathing but likely require further optimization including respiratory muscle strength training and cardiopulmonary rehabilitation strategies. For those patients, a cautious approach to ventilator weaning is adopted with a focus on cardiopulmonary optimization.

For patients with significantly impaired or absent diaphragm contractility, diaphragm atrophy becomes an important clinical feature (**Figure 4**). In general, an upper motor neuron injury that occurs above the anterior horn of the spinal cord results in impaired contractility of a muscle with minimal muscle atrophy in the subacute period when patients with new cSCI are admitted for inpatient rehabilitation. By contrast, lower motor neuron injuries occur due to damage to the anterior horn cell, the nerve root, or the peripheral nerve, and often result in severe muscle atrophy in the subacute period due to muscle denervation.^17^

**Figure 4.**
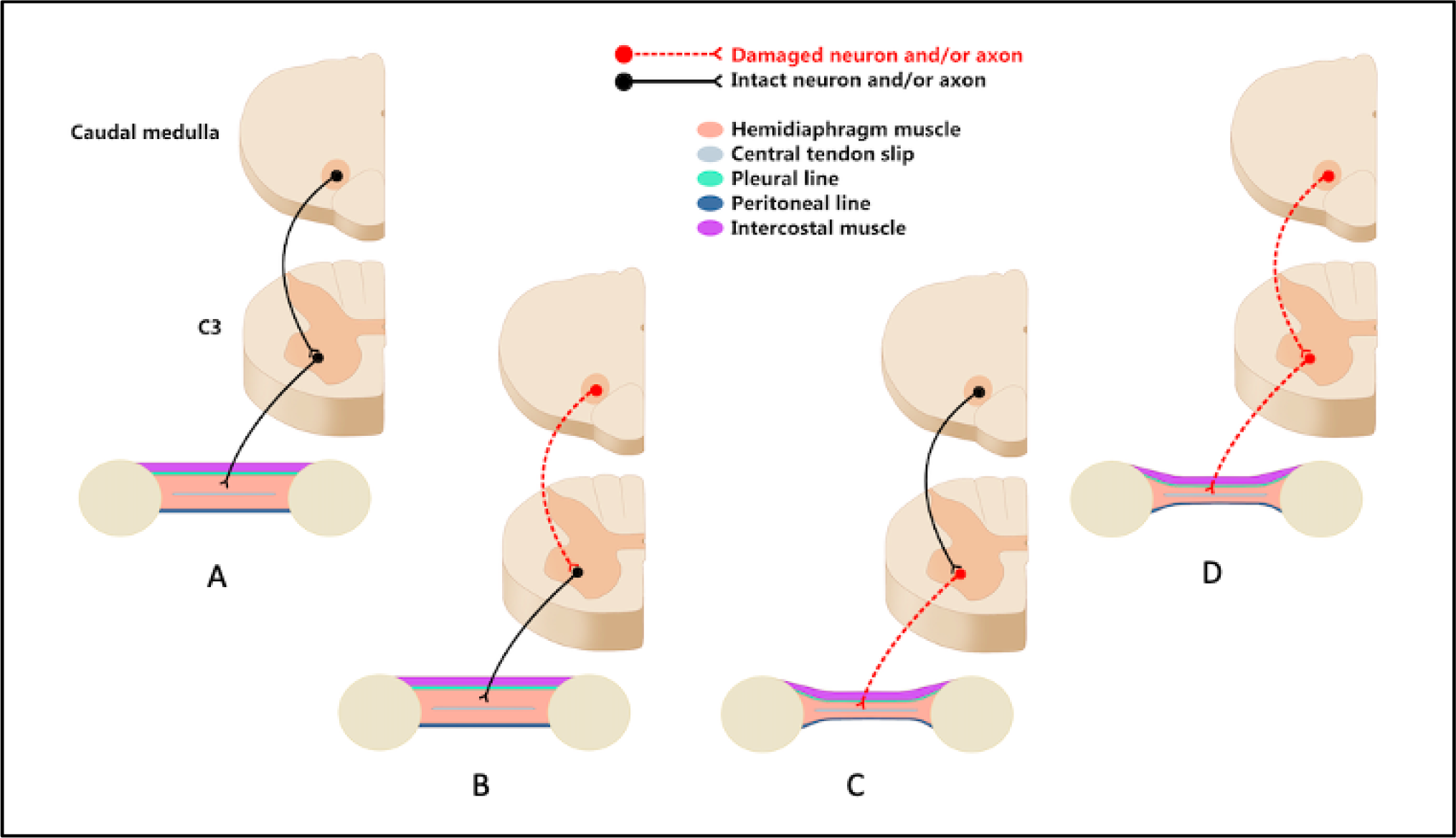
Illustration of upper vs lower motor neuron injury patterns at the C3 level of the spinal cord and peripheral nerve root. (A) Intact spinal cord and peripheral nerve, associated with a diaphragm with normal thickness. (B) Pure upper motor neuron spinal cord injury, which can result in impaired contractility of the diaphragm, but is associated with normal diaphragm thickness. (C) Lower motor neuron injury which can be associated with severe diaphragm atrophy due to axonal loss and denervation changes. (D) A mixed upper and lower motor neuron injury can result in severe diaphragm atrophy.

Patients who demonstrate absent contractility but normal thickness of the diaphragm (i.e. no atrophy) are identified as potential candidates for diaphragm muscle pacing, which involves implantation of electrodes to either the phrenic nerves at the neck or at the phrenic motor points within the diaphragm muscle itself. Both techniques require intact phrenic nerves, and denervation of the diaphragm is a contraindication. The benefits of diaphragm muscle pacing include partial or complete ventilator-free breathing, reduction in pulmonary infections, and improved comfort and speech production.^18–21^ At our center, a thoracic surgery team performs this evaluation and implantation of the pacer system.

Findings of lack of diaphragm contractility in conjunction with significant atrophy indicate denervation atrophy and axonal loss. In these cases, diaphragm pacing is not indicated. Other surgical interventions, such as nerve transfers, may be a possibility, and were recently considered in a prospective case series by Kaufman and colleagues.^22^ Additional studies are needed to further investigate the role of surgical interventions in achieving ventilator-free breathing after cSCI.

Limitations of our study include the retrospective study design and the single center setting. Though our sample size is relatively small, the patient population in question (ventilator-dependent patients with cSCI) is itself small in size, and our study represents a unique case series investigating the use of diaphragm ultrasound in this patient population.

Our data suggests that diaphragm ultrasound is a highly promising tool in prognosticating the ability to wean from a ventilator after cSCI. Future prospective studies are needed to better assess the impact of diaphragm muscle ultrasound as a tool to predict ventilator weaning outcomes.

## Data Availability

All data produced in the present study are available upon reasonable request to the authors

## 5. Acknowledgements

CKF would like to acknowledge support from the Belle Carnell Regenerative Neurorehabilitation Fund. SK would like to acknowledge support from the Catalyst Grant Program at Shirley Ryan AbilityLab. The authors acknowledge Dom D’Andrea for artwork used in Figure 4. The authors declare that this research was conducted in the absence of any commercial or financial relationships that could be construed as a potential conflict of interest.

---

**Supplemental Figure 1.**
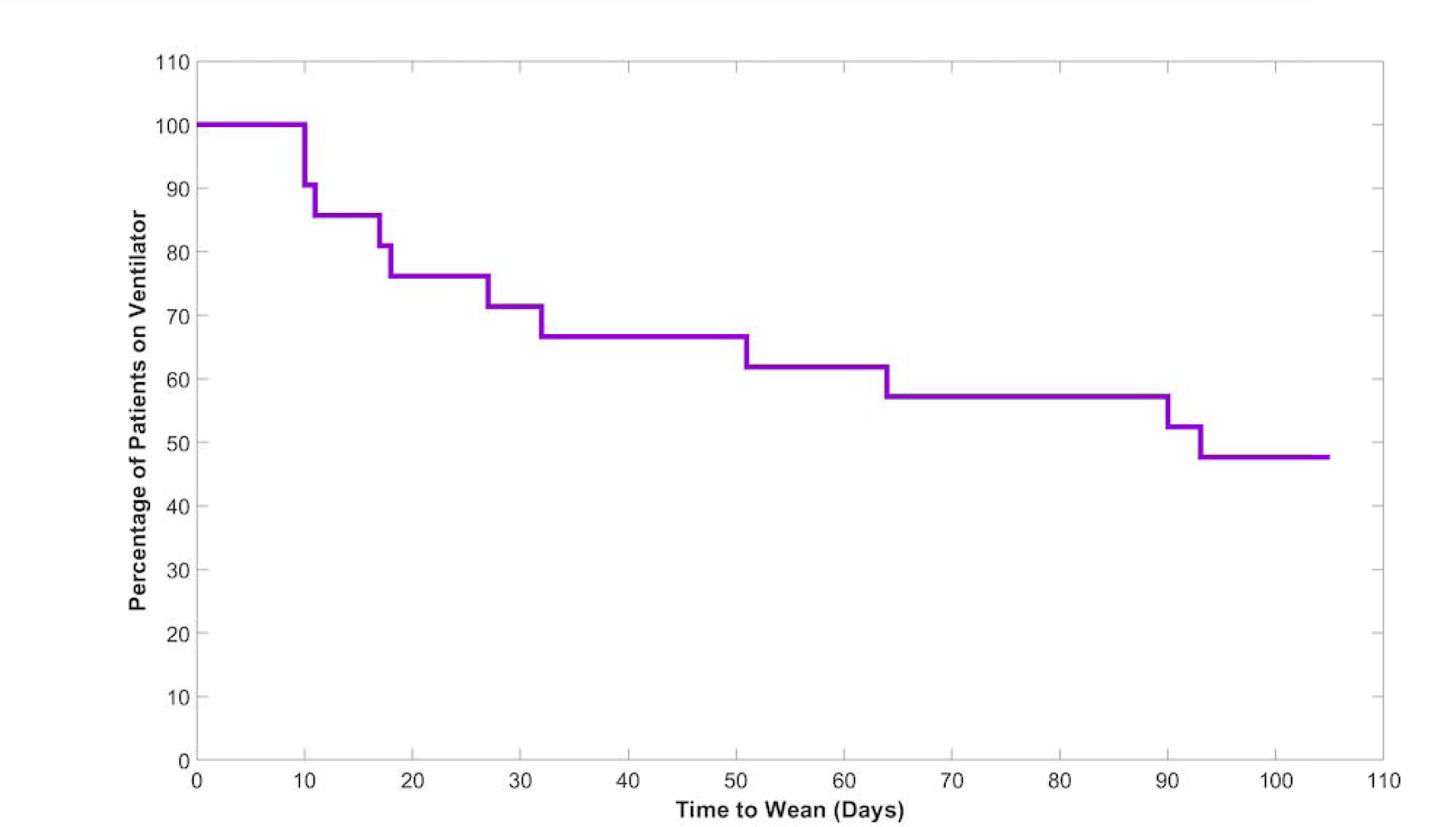
Kaplan-Meier curve demonstrating the probability of successful ventilator weaning as a function of length of weaning process.

**Supplemental Figure 2.**
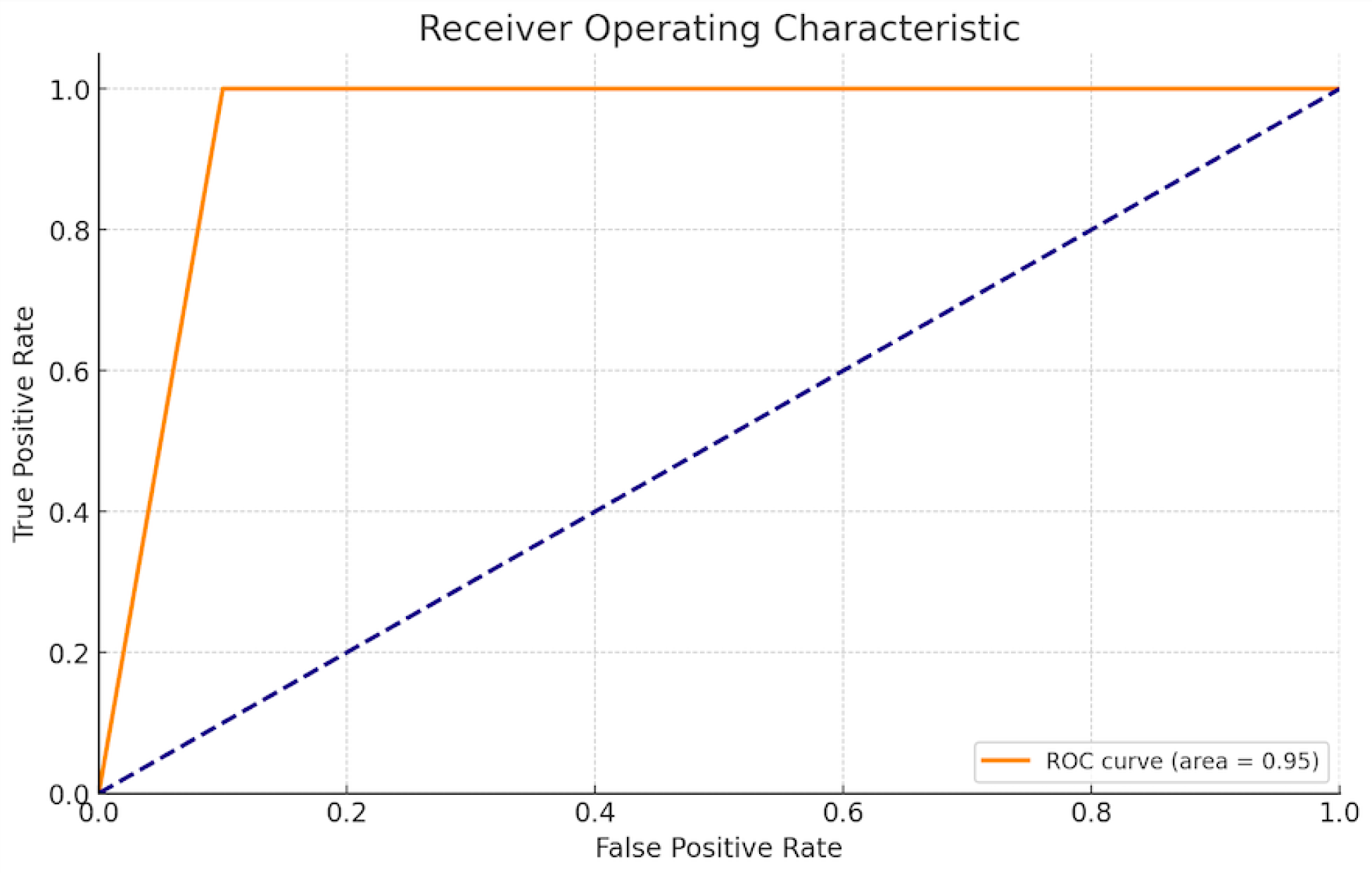
The Receiver Operating Characteristic (ROC) curve has been plotted with an area under the curve (AUC) of 0.95, which indicates a high overall performance of the best thickening ratio as a diagnostic test for successful ventilator weaning at the chosen cutoff of 1.2. The sensitivity (true positive rate) at this cutoff is 100%, meaning that all successful weaning cases were correctly identified using this cutoff. The specificity (true negative rate) is 90%, indicating that 90% of the failures were correctly identified as such when using the cutoff of 1.2. A high sensitivity and specificity suggest that a cutoff of 1.2 for the best thickening ratio is a good threshold for predicting successful ventilator weaning.

