## Supplemental File 1 for "The Predictive Value of Diaphragm Muscle Ultrasound for Ventilator Weaning Outcomes after Cervical Spinal Cord Injury: A Retrospective Case Series"

**Ventilator Weaning Protocol Summary**

**Initial Evaluation** for appropriateness of initial off ventilator trial

Vital capacity, negative inspiratory force, arterial blood gas, end tidal CO2.

Airway and tracheostomy tube clinical assessment.

Diaphragm ultrasound evaluation.

- **Initial Off Ventilator Trial** if patient meets inclusion criteria
  - Trial off ventilator with trach collar for 15 minutes while monitoring oximetry and vital signs every 5 minutes.

**Off Ventilator Progression**

Continue trach collar trials, increasing duration off the ventilator by 15-30 minutes every 48 hours until 3 continuous hours are tolerated.

Extend duration by 1 hour every 48 hours after reaching 3 hours, up to 12 continuous hours.

**Overnight and Final Assessment**

If all-day off ventilator is tolerated for 1 week, proceed to overnight off ventilator with continuous oximetry.

Assess for sleep-disordered breathing and consider non-invasive ventilation if oxygen desaturation index > 30 or SpO2 < 88% for more than 5-10 minutes.

Discontinue ventilator when continuous off ventilator trial is tolerated for 1 week.

**If Initial Trial Fails**

Continue to optimize cardiopulmonary status. Repeat trial in 2 weeks.

Explore alternative techniques, such as diaphragmatic pacing.
